# A comparative study and application of modified SIR and Logistic models at Municipal Corporation level database of CoViD-19 in India

**DOI:** 10.1101/2020.09.12.20193375

**Authors:** Naman S. Bajaj, Sujit S. Pardeshi, Abhishek D. Patange, Hrushikesh S. Khade, K. K. Mate

**Author notes:** Corresponding Author: *Abhishek D. Patange, Email ID.

## Abstract

The WHO declared a global pandemic owing to the newfound coronavirus, or Covid-19, in March 2020. The disease quickly spread around the world by contagion, and the lack of an appropriate vaccine has led to limited social activities in every track of life. Several national and state-level studies conducted predict the course of the pandemic using machine learning algorithms, most common being the SIR and the Logistic models. However, it is unclear whether these models work for a controlled community like Municipal Corporation or not. With measures now being employed at Municipal levels in India, it only fits to conduct particular research to examine how these models perform at lower jurisdictions. This study provides concrete evidence to show the superiority of the modified SIR model over the Logistic model based on analysis. The models not only give accurate predictions for up to 14 days but can also be used to define and signify the practicality and effectiveness of the decisions taken by the authorities. This feature of the study allows us to justly say that the government action of Unlock 1.0 was not a wise decision considering the nature of the pandemic. This study hopes to help the authorities to take the proper actions to prevent any further aggravation of the spreading virus. In conclusion, Municipal corporations having control should make use of this study to make decisions and test their effectiveness, and more corporations should be empowered to benefit from this study.

## 1. Introduction

The WHO declared a global pandemic of the newfound coronavirus, also named as Covid-19, in March 2020. Originated in Wuhan, China, the virus compromises the host’s respiratory system. The virus has had a comparatively lower death rate as compared to previous pandemics. However, the outbreak has disrupted the way of life for many and hit the world as a whole. Covid-19 is contagious and spreads through one’s respiratory droplets by coughing or sneezing. Once infected, it takes about a week or two for symptoms to show up. These can include mild to high fever, weakness, dry coughing, and shortness of breath in severe cases. The lack of a fitting cure or vaccine has limited the social activities in every track of life. Social distancing, self-quarantine, wearing a face mask, and frequent use of sanitizers are practiced extensively for the mitigation and control of the pandemic [1].

Be it the field of science and technology, the health sector, corporate worlds, share markets, or the daily applications used for entertainment and navigation, machine learning has proved to be useful. Also, significant production of data has become possible due to remarkable advances in biotechnology and health sciences [2]. Researchers are coming up with ways to better the algorithms, and scholars are using these methods to make useful predictions such as declaring a tumor fatal, treatability of cancer, to name a few. Pandemics are no exception to this, and machine learning algorithms are used to predict the turn of events.

The Susceptible-Infected-Recovered (SIR) model is an effective way of fitting data of the current pandemic. Several research studies were conducted using the same model and have proven to be beneficial. The SIR model is a compartmental model for modeling how a disease spreads through a population [3]. Another method is the logistic plots. A logistic curve fits the data of population growth efficiently. Data for the growth of the infected population exhibits the same nature, and hence logistic models are also used to predict pandemic states. Several studies comparing and proving these methods better than others or studies with adjusted SIR and logistic models are available publicly.

This particular study wishes to conclude with a comparative study between the SIR and logistic models. The primary goal is to present the results and help government organizations decide further steps needed to be taken to hinder the spread.

Section 2 takes the reader through the recent attempts to use SIR and logistic models for predictive analysis of the Covid-19 pandemic. The mathematics of the models is explained in Section 3. Section 4 details the case study and motivation regarding the same. The results of the models are presented in Section 5. Discussions and inferences are presented in Section 6, and Section 7 follow with the conclusions.

## 2. Literature review

As mentioned earlier, an extensive study was conducted on the coronavirus pandemic for various countries and states as well. The SIR model was the primary candidate for this. A simple SIR model for the UK and eight other European countries identified the pandemic characteristics as time-invariant across the world, and some highly variable, and also briefly studied the small but detectable average temperature effects on the probability α of infection [4]. Interestingly enough, the SIR model also predicts that high-risk individuals will be able to leave the lockdown well before vaccine arrival. The model was used to arrive at additional conclusions, presented as an exit time control problem where the lockdown ends with herd immunity [5]. The SIR model is compartmental. Also, many extensions of the same are proposed, in particular adding a compartment of exposed but non-contagious individuals, called the SEIR model [6] [7]. The bell-shaped distribution curves in the SIR model may not fit the Gaussian function as the CoViD-19 life cycle curves are not fully symmetric. A drawback of the SIR model is that it predicts the end dates to be much earlier than the actual end dates [8].

On the other hand, several scholars chose logistics modeling to predict the state of the pandemic. The efficacy of the model is a function of its fit to the available dataset. The question of whether we can find curve parameters that fit the complete dataset was addressed using a generalized logistic curve (or the Gompertz curve) for China and South Korea [9]. Another quantitative mathematical approach was able to derive a logistic model to characterize the age-specific case-fatality rates (CFRs). It was inferred that CFR does indeed increase with age [10]. A second peak was observed in the infections in late May of 2020 due to the relaxation of the mitigation efforts in the US. The basic logistic model fails to fit the data due to such relaxations, and an Adaptive Logistic Model (ALM) was designed, which gives more accurate predictions [11]. A Bayesian hierarchical five-parameter logistic model was fit to and approximated the observed data closely and could derive acceptable predictions [12]. Yet another logistic growth model was built for Italy, France, and Spain to help government authorities decide when to start the lockdown in a way not to exceed the health care capacities of the concerned country [13].

The search for a comparative study between the two models was short-lived for the lack of relevant literature. Forecasting of the spread of coronavirus in 8 different countries was undertaken based on both the logistic model and the SEIR and adjusted SEIR models [14]. An estimation of the final size of the pandemic for China, South Korea, and the rest of the world was done in two separate studies, one with the logistic model and the other with the SIR model [15]. Employing both models, the current stages of the pandemic in states of Karnataka, Kerala, and Maharashtra in India were predicted by a five-stage classification [16]. After realizing the existence of numerous variables at play at the Municipal level, the Indian Government allowed lower jurisdictions to take actions suitable to the concerned province. To explore the dynamics and nature of the pandemic in a controlled locality such as a Municipal Corporation, a comparative study was conducted that revealed exponential curves did not fit the Municipal data. Instead, a cubic curve gave better results, and more interestingly, the active cases followed a multi-peak Gaussian curve [17].

Gathering from above, several studies were conducted with the SIR and logistic models. Many tweaks were employed by scholars to achieve a better prediction from these models. However, there seems to be a lack of a detailed comparative study of these two methods, especially at a lower level. Till now, it is unclear whether these models work for a controlled community like Municipal Corporation. With measures now being employed at Municipal levels in India, it only fits to conduct particular research to examine how these models perform at lower jurisdictions. The aim is to help authorities take the correct steps based on these predictions to curb the mounting pandemic. With this motivation, this study intends to present the outcomes of both models at the National and Municipal level, by mapping and evaluating datasets of Akola, Mira-Bhayandar, Kalyan-Dombivli Municipal Corporation (KDMC), and India.

## 3. Mathematics of models

### 3.1 The SIR model

The compartmental nature of the model is seen in the following equations, where the total population is divided into three categories.

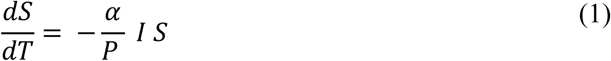

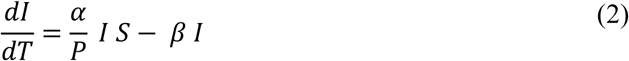

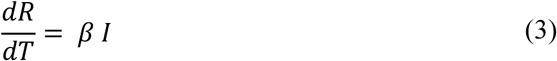

Here, at time T

*S*(*T*) = *S*: Number of susceptible people
*I*(*T*) = *I*: Number of Infected people
*R*(*T*) = *R*: Number of recovered people
*α*: Contact rate
1/*β*: Average infectious period

The total population size, P is then given as

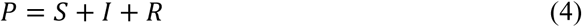

The initial conditions are taken as,

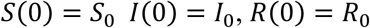

Solving equations (1) and (3) simultaneously and integrating, we get

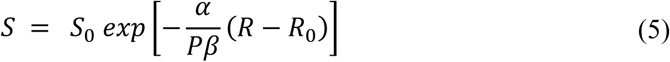

MATLAB’s *ode45* function was used to integrate the model equation.

Taking the limit as T →∞,

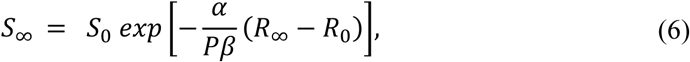

where *S*_∞_ is the number of susceptible people remaining and *R*_∞_ is the end number of recovered people. Here, the susceptible are taken to be the entire population and this is how it differs from the following modified SIR model.

### 3.2 Modified SIR model

The mathematics of the modified model is very much similar to that of the SIR model above and continued below. In the above model, the entire population is taken as the number of susceptible. However, younger people are found to be less susceptible to the virus. The COVID-19 pandemic has shown a markedly low proportion of cases among children where the age dependence in the probability of developing clinical symptoms rises from around 20% in under 10s to over 70% in older adults [18]. Also, people infected by the virus but are asymptomatic do not cough as frequently, which significantly reduces the chances of them spreading the virus. It was observed in 13 eligible studies that the symptomatic cases emerged to be a critical factor with very low transmission probability during the asymptomatic phase [19].

Hence, in equation (6), number of susceptible is not the entire population for the modified SIR model.

As there are zero infected people in the end, *I*_∞_ = 0 and from equation (4),

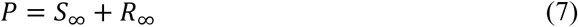

Plugging value of *S*_∞_ from equation (6),

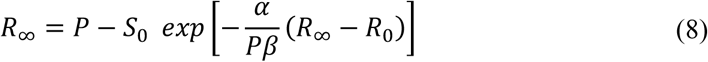

Number of recovered people at *T* = 0 *is R*_0_ = 0 and infected people, a constant *I*_0_ = *K*

We need to estimate the model parameters *α* and *β* and the initial values *S*_0_,*I*_0_. From the data, we can write the total number of cases *K* as

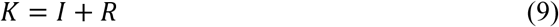

The estimated values of parameters and initial values give an estimate of *K* as 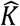 which we compare with the actual *K*.

The optimum values of the model parameters and the initial values can be found by minimizing the squared error or difference between the estimated and actual number of cases i.e.

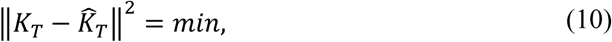

where,

*K_T_* = (*K*_1_,*K*_2_,*K*_3_,…,*K_n_*) are actual cases at times *T*_1_,*T*_2_,*T*_3_,…, *T_n_*

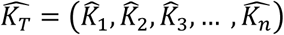 are respective estimates given by our model for a set of estimates of *α*, *β*, and initial values *S*_0_ and *I*_0_

The SIR model can be better grasped by observing the following flowchart.

**Figure 1:**
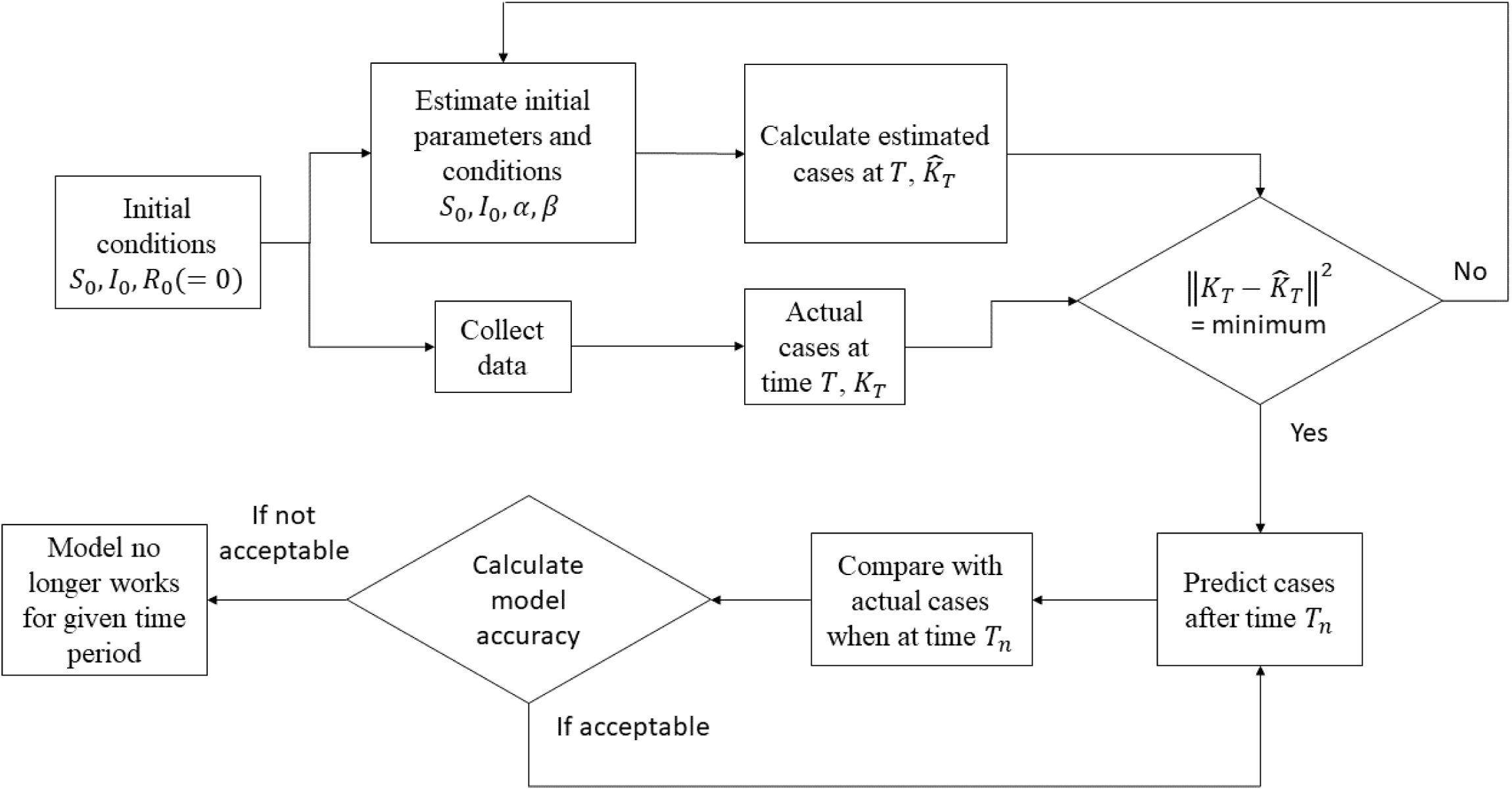
Flow diagram of SIR model

SIR and logistic models are often used to model epidemics as both are a good fit for the exponential growth rate [20]. As mentioned earlier, many scholars have used both models extensively, but separately to get appreciable results. Batista [15], in one of the few comparative studies found says, qualitatively, both models show that the epidemic is moderating for the time of February in case of China. The confidence level of a model tells us the range of possible deviation one might observe with respect to the real world data. Both SIR and logistic models give the narrowest confidence levels meaning their results are more reliable [20]. It therefore follows to study the mathematics of the logistic model to make a solid argument as to which model fits better for a Municipal Corporation level data of CoViD-19.

### 3.3 The Logistic model

The logistic growth model is as follows,

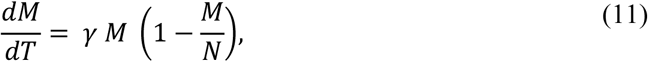

where

M – Accumulated number of cases
N – Final epidemic size (N > 0)
*γ* – Infection rate (*γ* > 0)

The solution for the non-zero positive initial number of cases is,

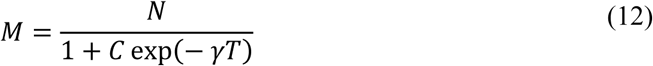

Here,

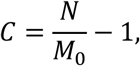

*M*(0) = *M*_0_

When T<<1, assuming final size is much larger than the initial cases, C>>1

We get natural growth,

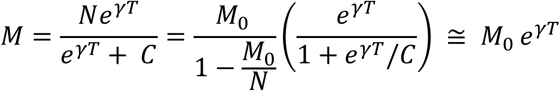

When T →∞, Weibull function is followed as,

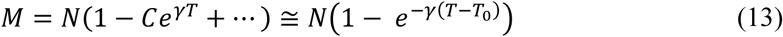

We reach the maxima for growth rate 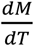 when 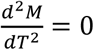

From this and above expressions of *M*, the growth rate peak is observed when

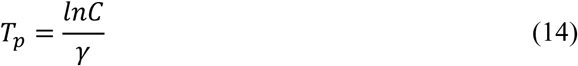

At this time *T_p_*,

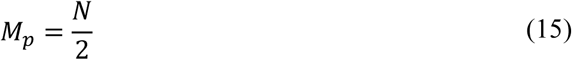

Also, the growth rate is,

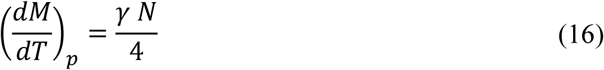

Often the intensity is judged by the doubling time, i.e., the time it will take to double the current number of the infected population.

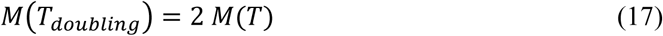

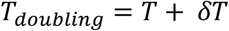

Solving for δ*T*,

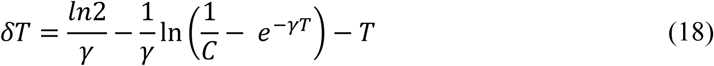

The first term represents initial exponential growth beyond which δ*T* increases with *T*.

As the infected population tends to the peak, doubling time tends towards infinity, i.e., when *M* → *N*/2,δ*T* →∞. Beyond *N*/2, doubling time is not defined as it represents the region beyond the peak.

One would want to know the final size of the pandemic *N* which can be estimated from all final size predictions *N*_1_, *N*_2_, *N*_3,_…, *N_n_* at times *T*_1_,*T*_2_,*T*_3,_…,*T_n_* respectively.

The iterated Shanks’ transformation gives

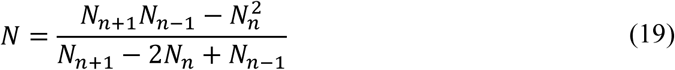

Here, care must be taken as there is no natural law behind the Shanks’ transformation, as in cases like *N*<*M_n_*, the calculated limit is useless. (*M*_1_,*M*_2_,*M*_3_,…,*M_n_*) are the total number of cases at the respective times)

The logistic model (12) is non-linear, and the initial guess should be provided carefully. As the equation (13) follows an exponential growth, predicting final size is difficult at the early stages. With enough data, however, one can use the following process to get a good initial condition.

Using *T* from equation (12) for 3 equidistant points

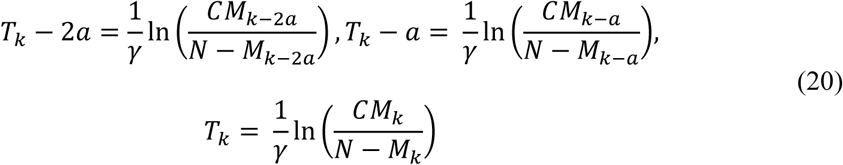

The following solution is acceptable if all unknowns are positive,

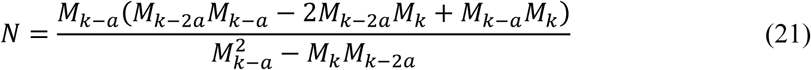

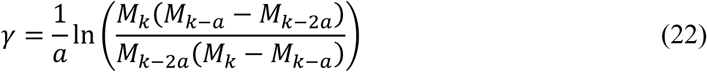

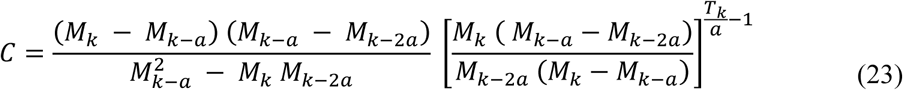

The above equations help to find an initial guess in the *fitVirus03* program. We choose the first, middle, and end data points for equations in (20). The regression analysis is questionable if this calculation fails. Finally, the parameters of the logistic model *N*,*γ*, and *A* are calculated by a least-square fit using the *lsqcurve* and *fitnlm* functions in MATLAB.

## 4. Methodology

The Indian subcontinent occupies most of South Asia and is ranked third in the list of worst-hit countries with over 2.4 million confirmed cases as of 16 August [21].

Data from 29 January to 9 June formed the database of the total reported and recovered cases, deaths, day-wise and active cases for a national-level analysis. Maharashtra stands as one of the most affected states in India, with over 844K confirmed cases as of 05 September [21].Three regions from Maharashtra: Akola, Kalyan-Dombivli Municipal Corporation (KDMC), and Mira-Bhayander were chosen for the statistical analysis. The official websites for the respective regions under scrutiny provided the data [22] [23] [24]. Akola’s statistics from 7 April to 9 June was computed. Data for Kalyan-Dombivli Municipal Corporation (KDMC) from 14 March to 9 June computed the results, whereas that of Mira-Bhayander was from 27 March to 9 June. The total cases, active patients, discharged patients, and deaths were listed in the data for each region for the respective time frames. The start date in each of the data was the date when they classified the first case.

The model predicted exponential and non-linear relations, which required sophisticated software to handle operations on data with ease. MATLAB was used for the statistical analysis of the above data and to plot it graphically.

## 5. Results

This section covers the results of SIR and Logistic models employed for each of the regions and India. The model parameters, statistics of the total cases and daily cases, and final states predicted by each of the three models are formulated in a table under every sub-section.

### 5.1 India

SIR model was used for two different timelines: data from start of pandemic to 30 May (Hereafter, SIR Model 1), and data from start of pandemic to 9 June (Hereafter, SIR Model 2) to incorporate the effects of Unlock 1 that took place on 1 June, 2020.

Figure 2 shows the prediction of the pandemic given by the SIR model 1 for the Indian population. The vertical red line in graph (2a) corresponds to the peak of daily new cases showcased by graph (2b). The total cases recorded every day are represented in a bar graph, and the growth rate is presented in percentage below. The news of Unlock 1.0 spread before 1 June and its effect on people was apparent since before 30 May. To highlight the difference between SIR 1 and SIR 2 clearly, data till 24 May was considered for India, as shown in Figure (2a).

**Figure 2:**
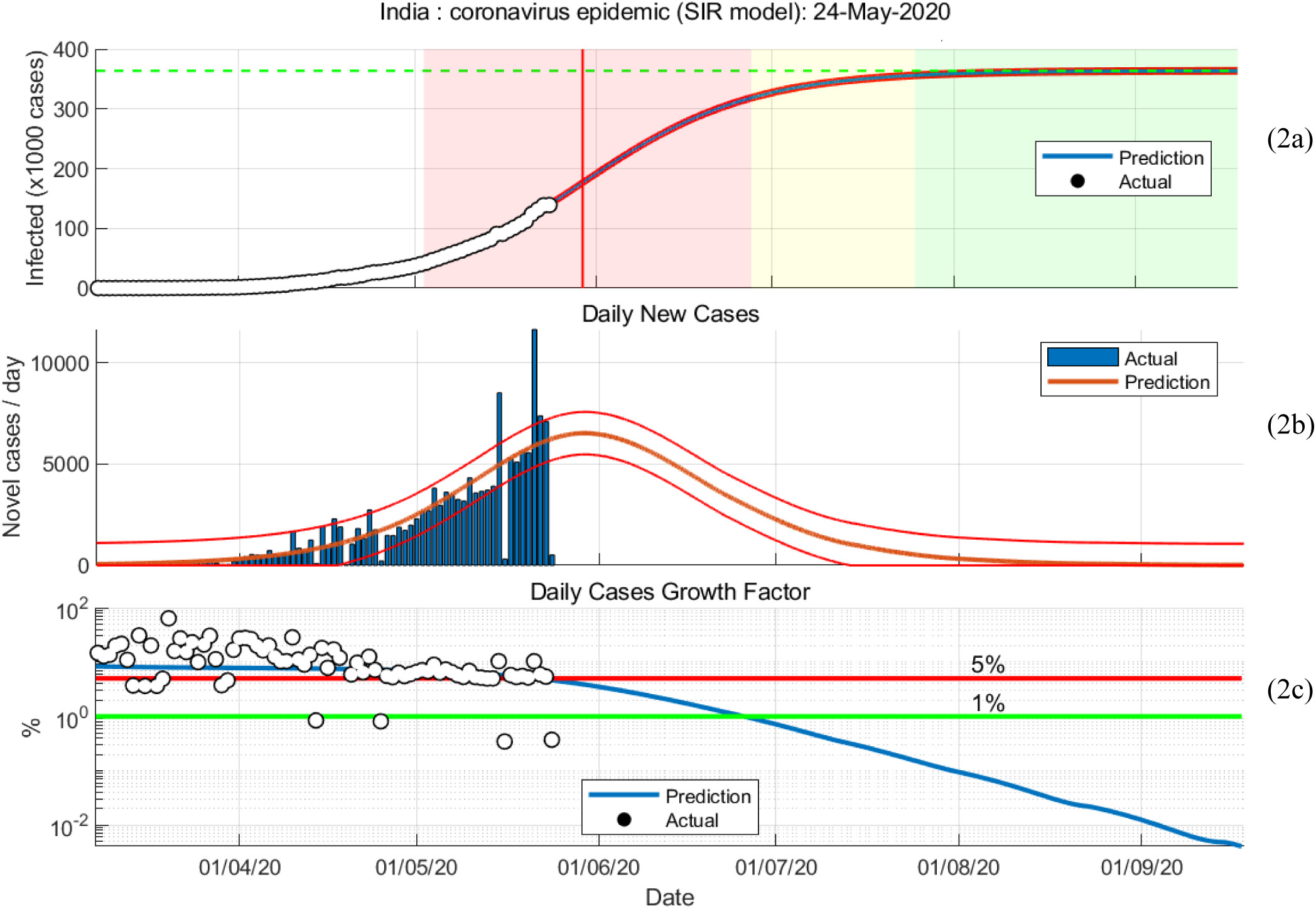
Graphical representations of SIR model 1 30-Jan to 24-May

The following set of graphs show the results of the SIR model 2, where the effect of unlock was included. Figure 3 shows the same in a model curve, the bar graph for daily cases where the peaks and their timeline is to be carefully noted, and the growth rate factor.

**Figure 3:**
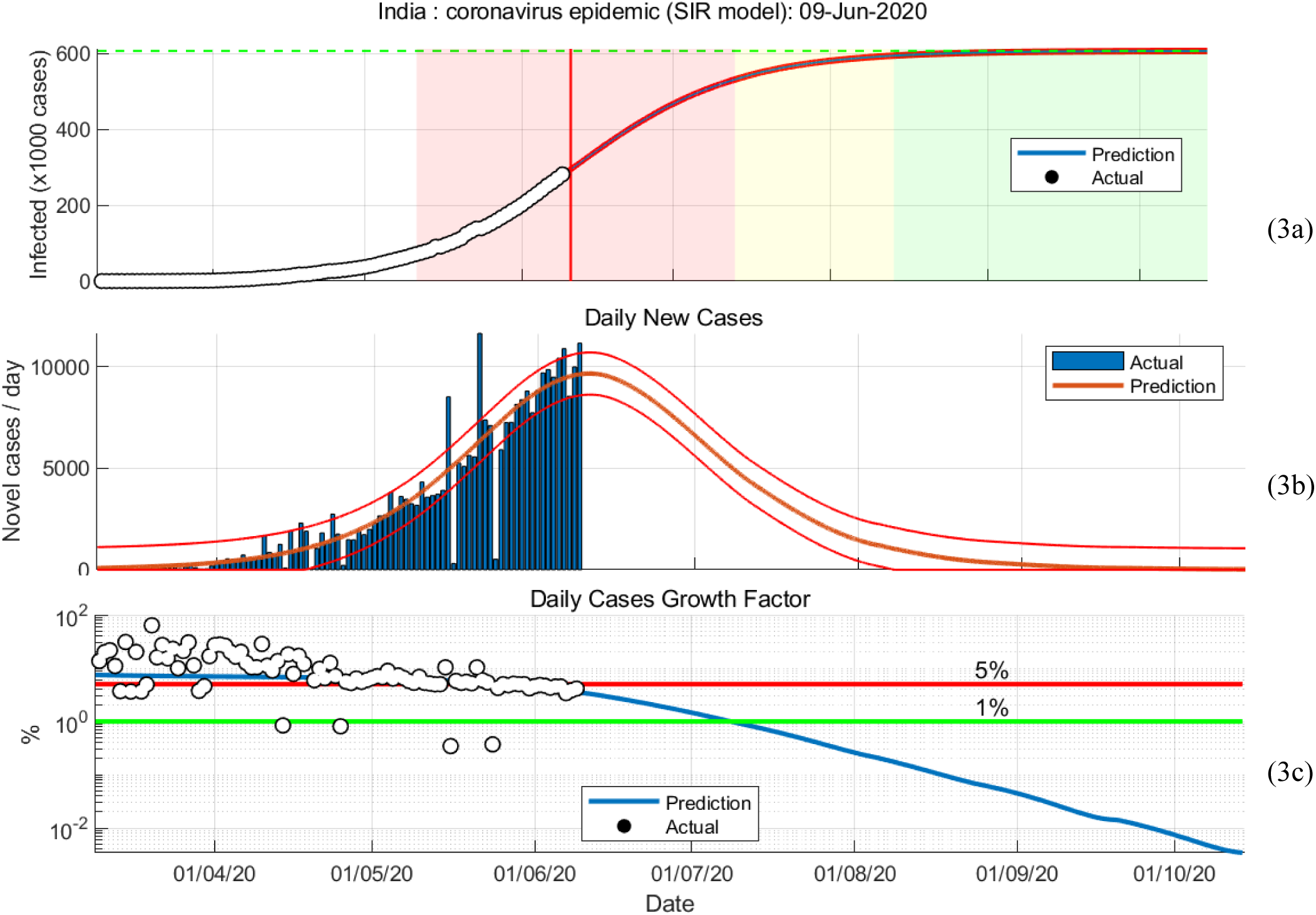
Graphical representations of SIR model 2 30-Jan to 09-Jun

Figure 4 shows the prediction given by the logistic model for India and the bar graph of daily cases.

**Figure 4:**
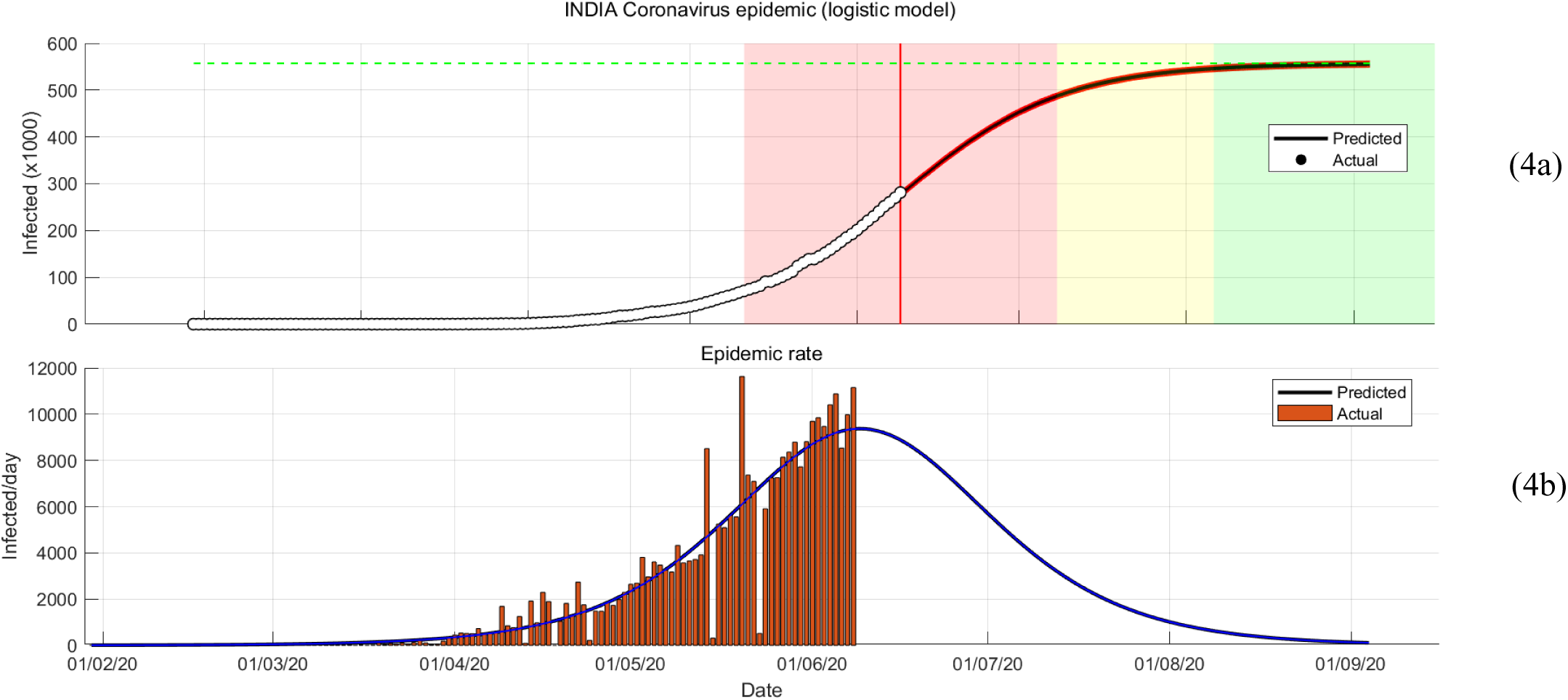
Graphical representations of logistic model 30-Jan to 09-Jun

**Table 1:** Parameters, statistical values and final state for India

| | Model parameters<br>(1/day) | | Root<br>mean<br>squared<br>error | $R^2$ | Number of<br>observations | p-value | Final<br>number<br>of cases | Final number<br>of susceptible<br>cases |
| --- | --- | --- | --- | --- | --- | --- | --- | --- |
| | $\alpha$ | $\beta$ | | | | | | |
| SIR<br>model 1 | 0.468 | 0.392 | 1561.99 | 0.998 | 114 | 4.20345e-149 | 363687 | 831729 |
| SIR<br>model 2 | 0.432 | 0.365 | 1913.81 | 0.999 | 130 | 1.81224e-198 | 606335 | 1.46633e+06 |
| | $N$ | $\gamma$ | | | | | | |
| Logistic<br>model | 277823 | 0.1257 | 1930 | 0.999 | 132 | 7.21e-214 | 557118 | - |

The table above formulates all significant statistical values and the model parameters for the models’ SIR 1, SIR 2, and logistic.

The difference between SIR models 1 and 2 is clear from Figures 1 and 2 plotted for India. The graphs for the same were plotted for regions below but not shown henceforth. The difference between the two models for the Municipal Corporations is apparent from the results in Table 5.

### 5.2 Akola

Figure 5 below shows the prediction by the SIR model 2. Peaks correspond to the dates of significant developments, and the graphs show a change in the daily number of cases.

**Figure 5:**
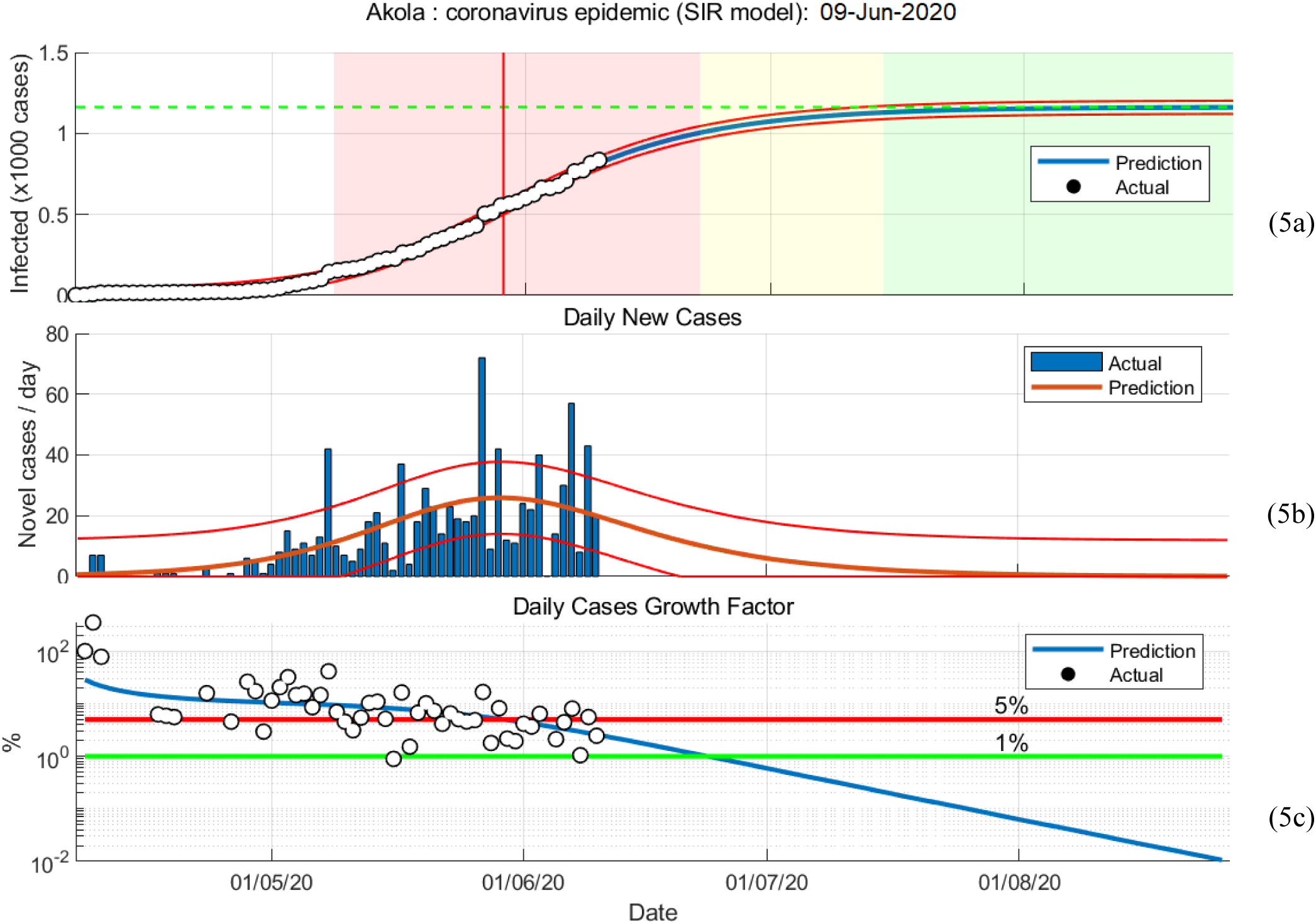
Graphical representations of SIR model 2 07-Apr to 09-Jun

Similarly, Figure 6 shows the pandemic for the same timeline but using the logistic model.

**Figure 6:**
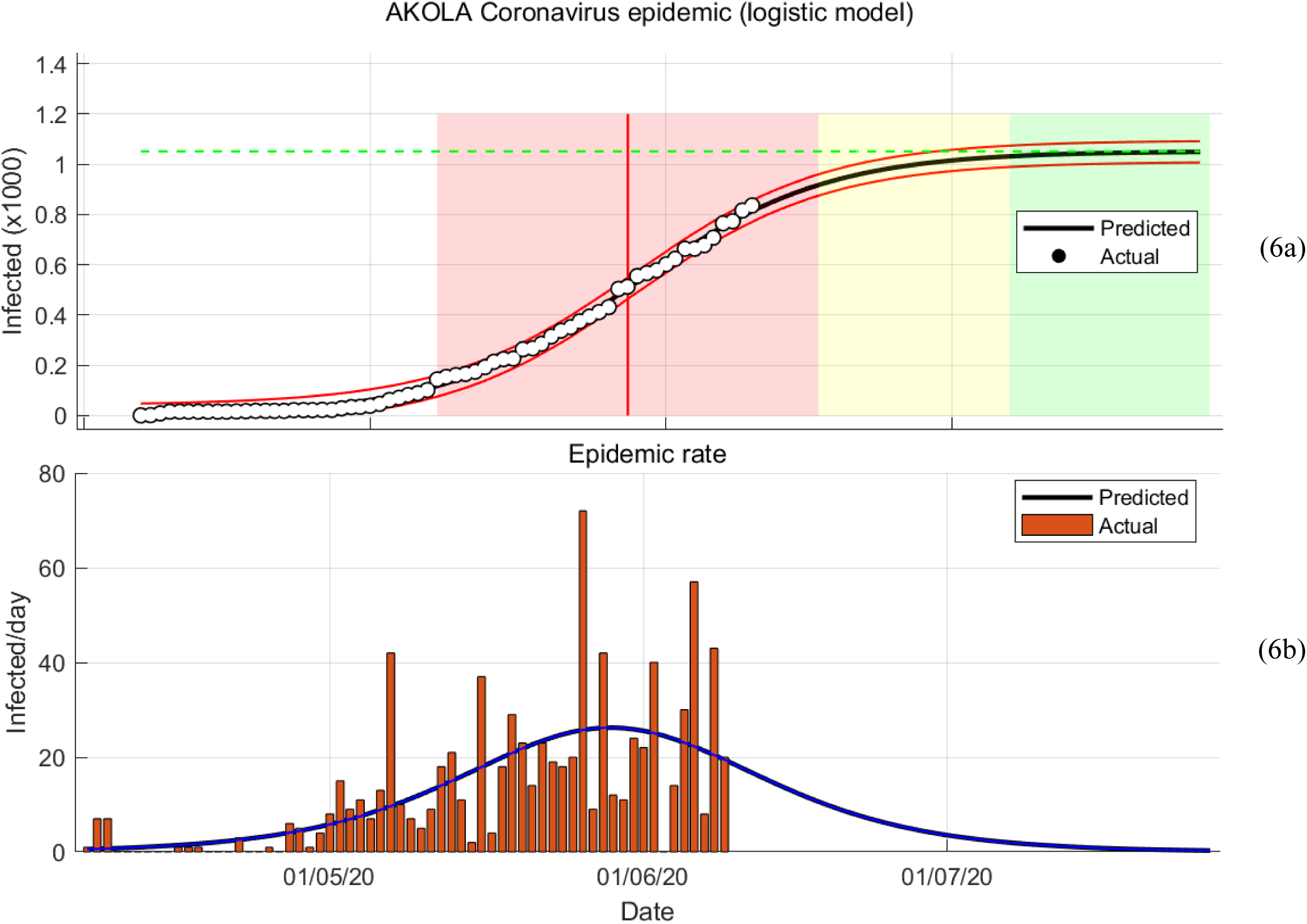
Graphical representations of logistic model 07-Apr to 09-Jun

The model parameters, accuracies and final predictions for Akola are put together in the table below, Table 2.

**Table 2:** Parameters, statistical values and final state for Akola

| | Model parameters<br>(1/day) | | Root<br>mean<br>squared<br>error | $R^2$ | Number of<br>observations | p-value | Final<br>number<br>of cases | Final number<br>of susceptible<br>cases |
| --- | --- | --- | --- | --- | --- | --- | --- | --- |
| | $\alpha$ | $\beta$ | | | | | | |
| SIR<br>model 1 | 0.184 | 0.077 | 12.3954 | 0.997 | 60 | 4.11848e-70 | 1040 | 143 |
| SIR<br>model 2 | 0.271 | 0.173 | 13.5604 | 0.997 | 65 | 9.7425e-79 | 1162 | 696 |
| | $N$ | $\gamma$ | | | | | | |
| Logistic<br>model | 856.33 | 0.16884 | 14 | 0.997 | 65 | 1.28e-87 | 1051 | - |

### 5.3 KDMC

The Kalyan-Dombivli Municipal Corporation’s official website provided the data for plotting the predictions of SIR model 2 and the logistic model, as shown in the following set of graphs.

The graph (7b) shows the variation of daily new cases in the form of a single peak Gaussian curve. Different parameters were chosen for the model to get a range of predictions to increase the accuracy of predictions. The use of Gaussian is also beneficial to detect any anomalous behavior in the trend of daily new cases, which can then be associated with specific policies employed or actions taken around that timeline.

**Figure 7:**
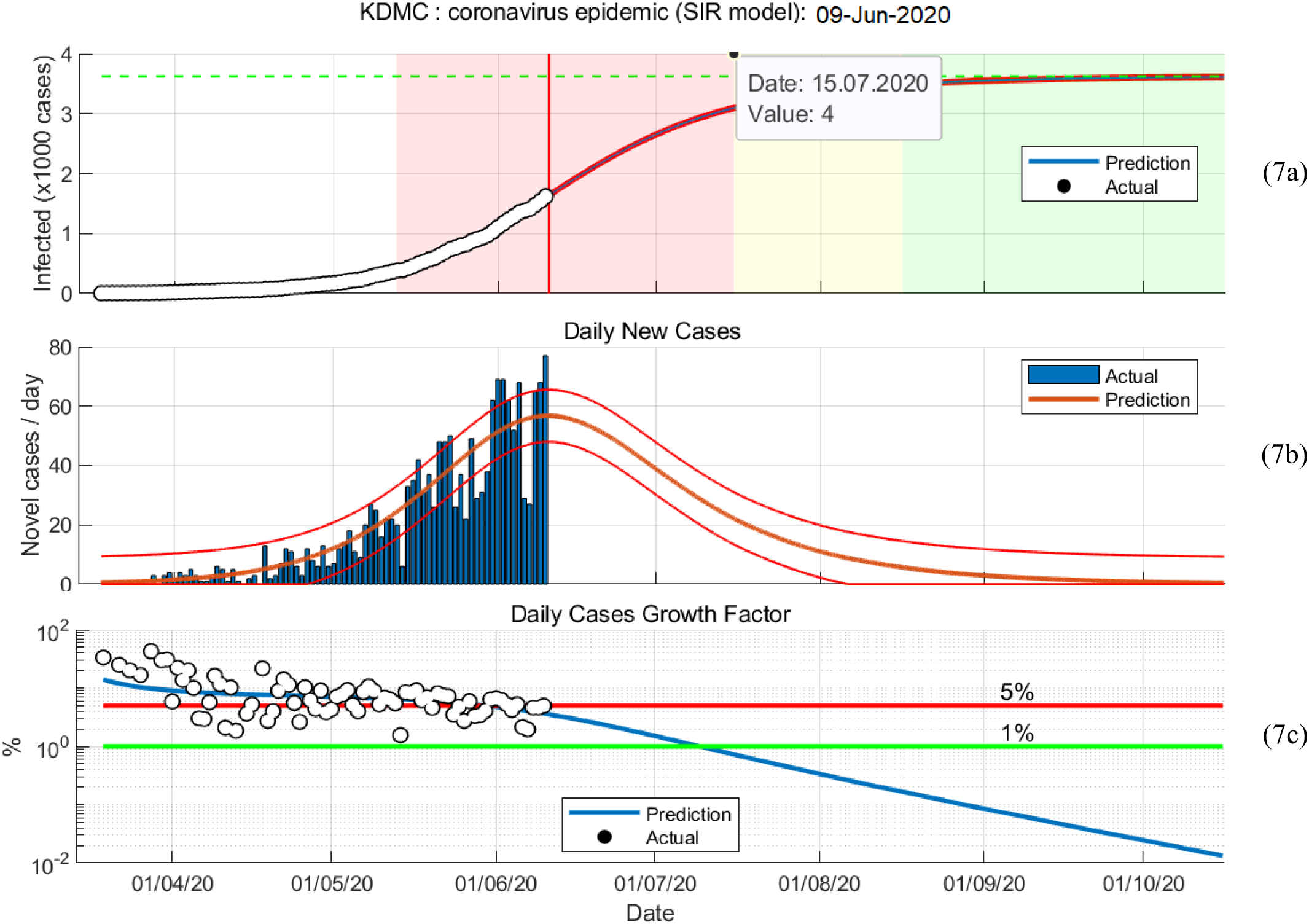
Graphical representations of SIR model 2 14-Mar to 09-Jun

A logistic model employed for the data of KDMC in the same timeline shows the trend represented by Figure 8.

**Figure 8:**
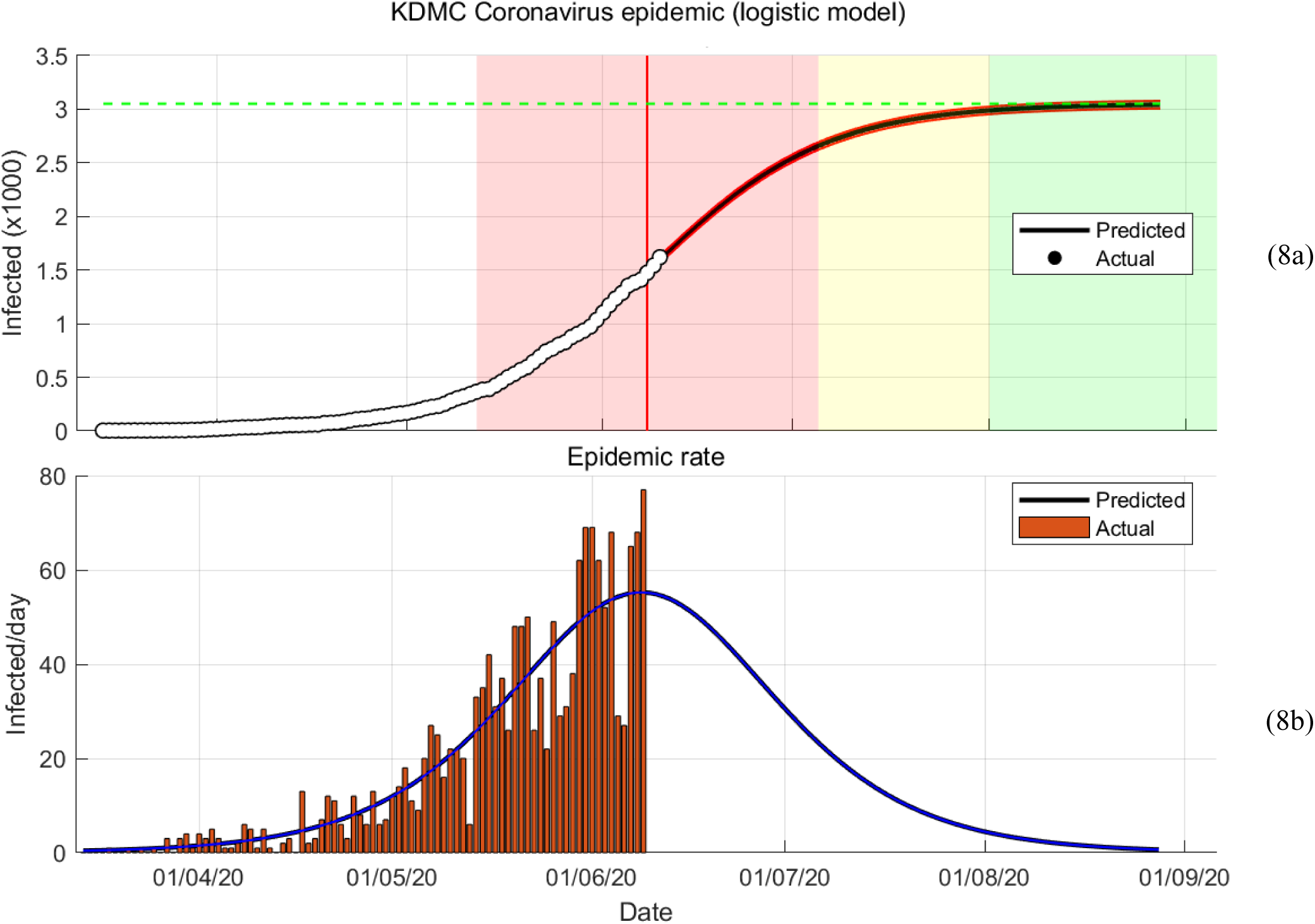
Graphical representations of logistic model 14-Mar to 09-Jun

As listed for the previous region and India, Table 3 above summarizes the details of the model employed, its results along with the statistical data, including accuracy, for KDMC.

**Table 3:** Parameters, statistical values and final state for KDMC

| | Model parameters<br>(1/day) | | Root<br>mean<br>squared<br>error | $R^2$ | Number of<br>observations | p-value | Final<br>number<br>of cases | Final number<br>of susceptible<br>cases |
| --- | --- | --- | --- | --- | --- | --- | --- | --- |
| | $\alpha$ | $\beta$ | | | | | | |
| SIR<br>model 1 | 0.09 | 0.014 | 9.8400 | 0.999 | 74 | 3.91967e-102 | 2487 | 3 |
| SIR<br>model 2 | 0.135 | 0.064 | 11.9907 | 0.999 | 85 | 1.77192e-128 | 3625 | 756 |
| | $N$ | $\gamma$ | | | | | | |
| Logistic<br>model | 2096.1 | 0.0904 | 11.7 | 0.999 | 89 | 7.13e-146 | 3051 | - |

### 5.4 Mira-Bhayander

Results of the Mira-Bhayander region include the depiction of the employed models, i.e., SIR models, with and without consideration of effects of unlock on the behavior of the masses, and the logistic model.

The growth factor features the growth in the daily cases relative to the previous day. This is showcased in the graph (9c), where the growth rate is almost constant, over 5%, and then reduces significantly after the bell curve passes the peak. The graph below shows the logistic model for the Mira-Bhayander and the variation of daily cases with time followed by the summary of results in Table 4.

**Figure 9:**
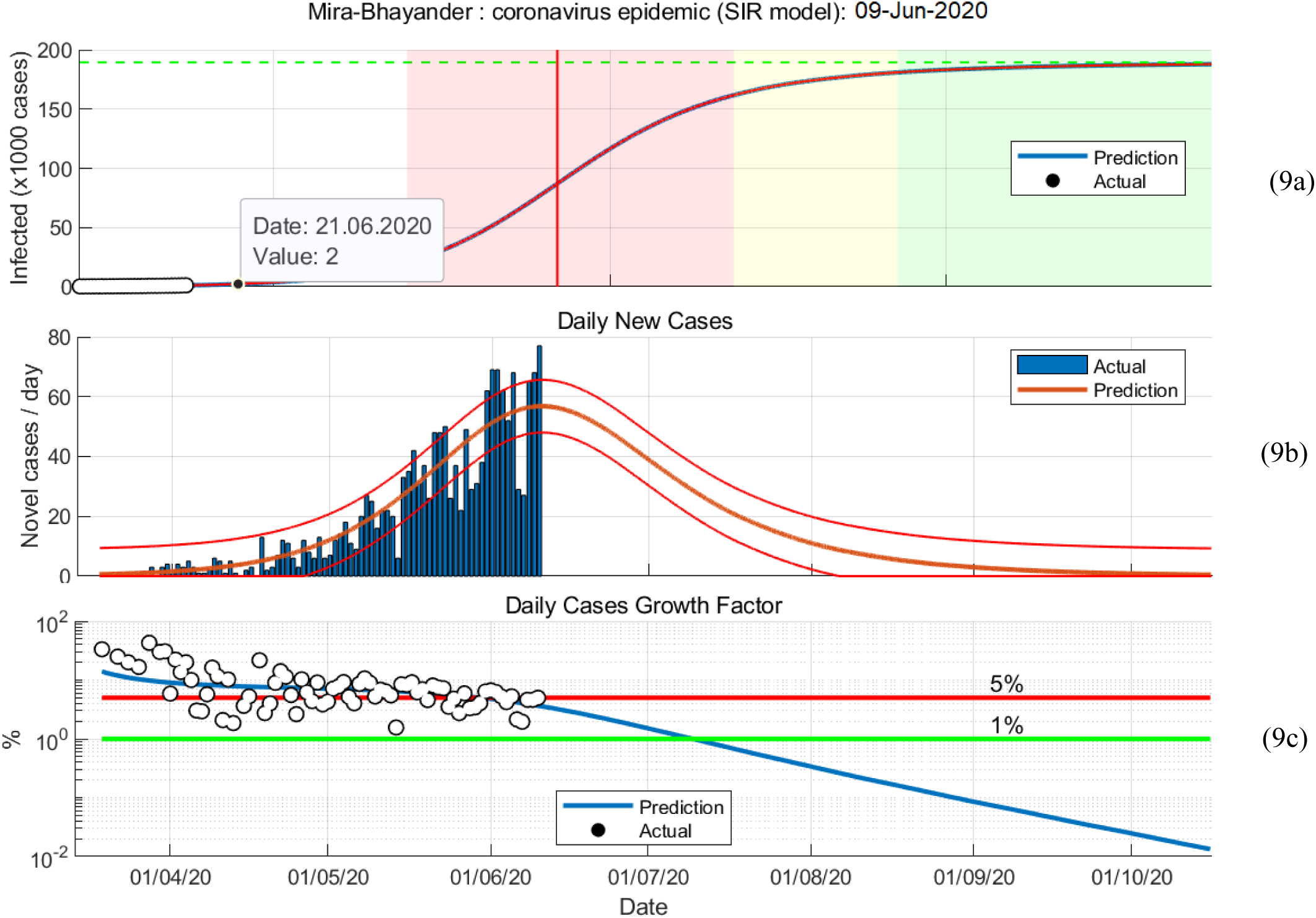
Graphical representations of SIR model 2 27-Mar to 09-Jun

**Figure 10:**
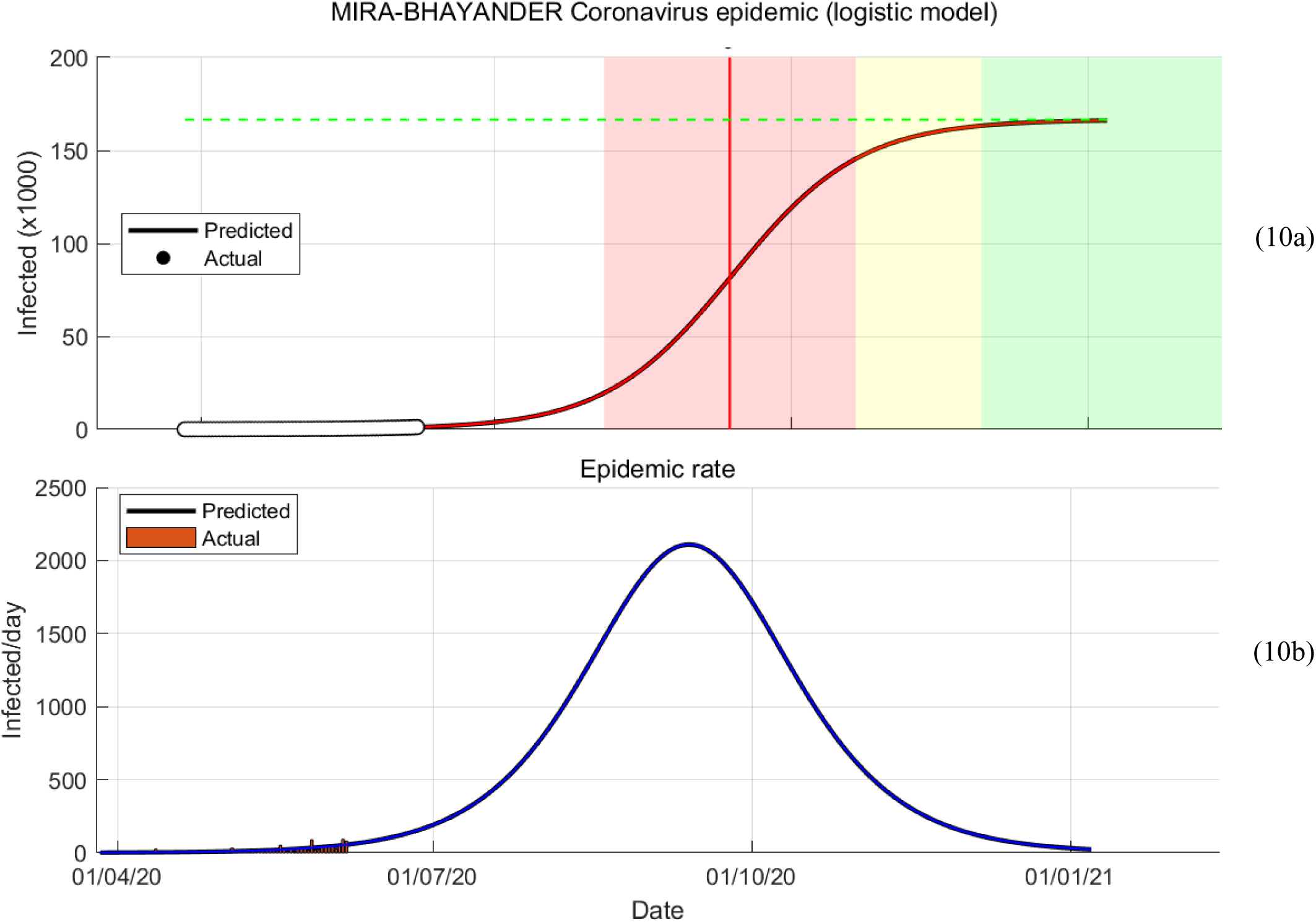
Graphical representations of logistic model 27-Mar to 09-Jun

**Table 4:** Parameters, statistical values and final state for Mira-Bhayander

| | Model parameters<br>(1/day) | | Root<br>mean<br>squared<br>error | $R^2$ | Number of<br>observations | p-value | Final<br>number<br>of cases | Final number<br>of susceptible<br>cases |
| --- | --- | --- | --- | --- | --- | --- | --- | --- |
| | $\alpha$ | $\beta$ | | | | | | |
| SIR<br>model 1 | 0.068 | 0.019 | 16.1238 | 0.994 | 65 | 1.78123e-68 | 43105 | 1334 |
| SIR<br>model 2 | 0.065 | 0.016 | 16.3196 | 0.997 | 73 | 1.1228e-88 | 189464 | 3195 |
| | $N$ | $\gamma$ | | | | | | |
| Logistic<br>model | 1221.8 | 0.1547 | 16.3 | 0.997 | 73 | 4.45e-105 | 166576 | - |

All tables are to be read carefully to understand the difference between the results given by the SIR models and the logistic model at the national and the regional level.

## 6. Inferences

- The modified SIR models, namely, SIR 1 and SIR 2 were employed for COVID-19 cases till 24 May and 9 June respectively. The predicted cases of SIR 2 model were approximately 60% more than the total cases predicted by SIR 1 model. This extreme and significant deviation in results can be attributed to the fact that SIR 2 incorporates the effect of Unlock 1.0, introduced on 1 June 2020, after the outbreak of COVID-19, whereas SIR 1 doesn’t.
- Table 5 depicts a comparison between the actual cases on 23 June 2020 versus the predicted results obtained after employing modified SIR 1, modified SIR 2 and logistic model on their respective datasets. Both SIR 2 and logistic model take into account the impact of Unlock 1.0, thus giving higher accuracies than SIR 1 model. Furthermore, the superiority of modified SIR 2 model over the Logistic model can be concluded on the basis accuracy of the results obtained.
- The actual cases obtained for Mira-Bhayandar even on 5 July match the results obtained from modified SIR 2 model with 90% accuracy i.e. 25 days after analysis.
- For districts or municipalities showing fairly lesser number of cases, for example Akola, the model starts to fail due to the slow progress of the pandemic. Though for Akola, the accuracy is still 80%.
- The sixth degree polynomial regression models will help Indian doctors and the Government in preparing their plans for the next 7 days [25]. According to the study conducted by Bajaj et al. [17], it only fits to consider that the prediction accuracy for Municipal Corporation level would decrease due to an increase in parameters under consideration. However, not only does the model suggested by this study give high accuracy but it also provides prediction up to 14 days, i.e. double the time period of the study mentioned above.

**Table 5:** Accuracy of models, 23 June 2020

|  | Actual cases | Modified SIR 1 |  | Modified SIR 2 |  | Logistic |  |
| --- | --- | --- | --- | --- | --- | --- | --- |
|  |  | Predicted cases | Accuracy | Predicted cases | Accuracy | Predicted cases | Accuracy |
| India | 449394 | 315000 | 70.09 | 430000 | **95.68** | 405000 | 90.12 |
| Akola | 1272 | 1030 | 80.98 | 1060 | **83.33** | 1024 | 80.50 |
| KDMC | 2541 | 2050 | 80.68 | 2400 | **94.45** | 2300 | 90.52 |
| Mira-Bhayander | 2481 | 2440 | 98.35 | 2590 | **99.64** | 2500 | 99.23 |

## 7. Conclusions

- From the rise in cases observed and predicted by modified SIR models 1 and 2, it can justly be said that the introduction of Unlock 1.0 was not a wise decision considering the nature of pandemic.
- Modified SIR 2 model is far better than Logistic model as it provides prediction result more accurately. Even though this model has previously only been employed to predict the outbreak statistics for larger areas like countries or states, it can be unequivocally used for Municipal Corporations such as Mira-Bhayandar or localities with slow growth of the pandemic. Furthermore, if no significant environmental changes are introduced in controlled areas like Mira-Bhayander, it can provide prediction accuracy of 90% for at least one and at most 2 months rather than previously considered 14 days period.
- Analysis of Akola dataset set shows that the accuracy of the model reduces significantly (only 80%) for areas with cases lower than 1000.
- Even though the model cannot be utilized for longer time periods, but a remarkable method to use it would involve predicting the number of cases from the day any new policy to combat COVID-19 is introduced and thereby recommending periodic review and analysis. The results obtained from both analysis would define and signify the practicality and effectiveness of the decision.
- Further, regions similar to each other in terms of population, location, and available facilities like manpower, can then implement the same decisions which proved beneficial and can help curb the pandemic in all such regions.
- Few states in India like Maharashtra have already extended the authority to take required actions against the increasing pandemic to Municipalities. This study will help these government offices to take steps in the right direction. The author recommends corporations that have control to use this study as a tool and take the right decisions. It is also recommended to extend authority to more Municipal Corporations and benefit from the model.

## Data Availability

All datasets were acquired from official websites of respective location

https://www.covid19india.org/

https://www.mbmc.gov.in/master_c/important_information

https://dashboard.kerala.gov.in/

https://dio-akola.blogspot.com/

